# Medical Risk Classification For Severe COVID-19 Based On Chronic Medical Conditions: A Comparative Analysis

**DOI:** 10.1101/2024.09.06.24313189

**Authors:** Ilse Westerhof, Annemarijn de Boer, Angela Lupattelli, Isabel Slurink, Nicolaas P.A. Zuithoff, Otilia Boldea, Hedvig Marie Egeland Nordeng, Jizzo R. Bosdriesz, Frank Pijpers, Maarten Schim van der Loeff, Mirjam Knol, Janneke van de Wijgert, Patricia Bruijning-Verhagen, Ganna Rozhnova

## Abstract

**Background:** The European Centre for Disease Prevention and Control lists medical conditions conferring high- or moderate-risk for severe COVID-19. Individual European countries also developed risk classifications to guide COVID-19 vaccination recommendations.

**Objectives:** To examine discordance between national and European risk classifications for severe COVID-19 in the Netherlands and Norway across general, hospitalized, and deceased populations, and investigate related factors.

**Methods:** This multi-country data-linkage study included 17.4 million inhabitants in the Netherlands and 5.6 million in Norway in 2020. Medical conditions were defined using ICD-10 codes in the European and Dutch classifications, and ICD-10 and ICPC-2 codes in the Norwegian classification. Individuals were classified as high-, moderate-, or low-risk per each classification. Performance was evaluated using sensitivity, specificity, positive and negative predictive values (PPV, NPV), Brier score, and area under the curve (AUC).

**Results:** Discordance between classifications was 12–14% in the general population and 36-47% among hospitalized or deceased individuals. The European classification assigned more individuals to high-risk, while national classifications assigned more as moderate-risk. National classifications demonstrated higher sensitivity, particularly in Norway, while the European classification showed higher specificity. PPVs were ≤0.9% and NPVs near 100% across all classifications. Norway’s national classification had better discrimination and calibration, while in the Netherlands, the European classification better predicted mortality. Discordances frequently involved subjects with cardiovascular disease, lung disease, and diabetes.

**Conclusion:** The European classification defines a substantially broader high-risk population compared to national classifications. No single classification is universally optimal. Further research should refine which chronic conditions best predict severe COVID-19.

**Funding:** ZonMw, EU, FCT, Norwegian Research Council’s COVID-19 Emergency Call, iAPOGEE.

## Introduction

The impact of public health measures, including COVID-19 vaccination, depends on the accurate identification of populations at elevated risk for severe outcomes, including hospitalization and death [1]. The World Health Organization recommends targeting vaccination programs to population groups at the highest risk of severe COVID-19 [1], but disparities exist in the risk classifications developed by different health authorities. While COVID-19 currently presents as common cold- or flu-like illness, older individuals and those with certain chronic medical conditions still face higher hospitalization and death rates compared to the general population [2,3]. Most risk classifications group individuals into high-, moderate-, and low-risk for severe COVID-19, based on factors like medical conditions and age. High-risk individuals have chronic conditions that lead to a much higher likelihood of severe outcomes, while those in the moderate-risk group face an elevated but lower risk.

Informed by influenza risk classifications and studies on risk factors for severe COVID-19, national (e.g., the Health Council of the Netherlands[4] and the Public Health Institute in Norway [5]) and international (e.g., the European Centre for Disease Prevention and Control [6]) public health authorities assigned high- or moderate-risk of developing severe COVID-19 to specific pre-existing medical conditions. These medical risk classifications have remained unchanged [6,7] or undergone minor alterations [4,5,8,9] since their development at the beginning of the pandemic. They have continued to guide epidemiological research and policy decisions on eligibility criteria and prioritization for vaccination programs aimed at reducing the incidence of severe COVID-19. While these international and national classifications can be applied within the same country, it is unclear to what extent they differ in assigning medical risk status to individuals.

To inform evidence-based decision-making for severe COVID-19, we conducted a comparative analysis of risk status based on a national versus the European classification. This observational national registry-based cohort study used Dutch and Norwegian health and administrative data from 1 January 2020, linked at the individual level, including 17.4 million inhabitants of the Netherlands and 5.6 million inhabitants of Norway. We aimed to examine discordance between the respective national and European medical risk classifications for severe COVID-19 for the Netherlands and Norway across general, hospitalized, and deceased populations, and investigate related factors.

## Methods

At the start of the COVID-19 pandemic, the Netherlands and Norway adopted national vaccination strategies, with the shared objective of minimizing severe COVID-19 outcomes such as hospitalization and death, reducing pressure on the health system, and preventing social disruption [10]. Individuals with chronic medical conditions associated with increased risk of severe COVID-19 were prioritized early in the national vaccination campaigns, which began in January 2021. In the Netherlands, high-risk individuals were identified through the International Statistical Classification of Diseases and Related Health Problems (10^th^ revision; ICD-10) diagnostic codes [11]. The National Institute of Health and the Environment (RIVM) and Health Council defined the medical risk groups recommended for vaccination, including people with cardiovascular disease, diabetes, chronic lung or kidney disease, immunosuppression, cancer, and severe obesity [9,12]. The Norwegian Institute of Public Health (FHI) used both ICD-10 and International Classification of Primary Care (ICPC-2) [13] codes in the national health registries to define and flag individuals with moderate- or high-risk conditions for prioritized vaccination [5].

### Data Collection

#### Data collection – The Netherlands

The Dutch dataset included hospital discharge registry (DHD) and the National Cause of Death Registry (NCDR) data for all inhabitants of the Netherlands as per the Dutch Population Registry on 1 January 2020, as well as individuals hospitalized and deceased due to COVID-19 in 2020. The three registries were linked using unique personal identification numbers. The Population Registry was used to extract age, immigration, and emigration dates. The DHD provided information on chronic medical conditions diagnosed in hospitals since 1 January 1995, including the date of diagnosis. All diagnoses were registered according to the ICD-10 codes. From the NCDR, the date of death was extracted.

#### Data collection – Norway

The Norwegian dataset included data from multiple Norwegian registries for all inhabitants of Norway as per the National Population Registry on 1 January 2020, as well as individuals hospitalized and deceased due to COVID-19 in 2020. The registries included the Statistics Norway (SSB) registry, the Norwegian Patient Registry (NPR), and Norway Control and Payment of Health Reimbursements (KUHR) registry, and the Norwegian Cause of Death Registry (DAR). These registries were linked using unique personal identification numbers. The NPR and SSB registries were used to extract age, immigration, and emigration dates for all inhabitants of Norway. The KUHR registry provided information on medical diagnoses in primary and secondary outpatient care using ICPC-2 and ICD-10 codes, respectively. The NPR provided information on medical diagnoses in specialist outpatient and inpatient care, using ICD-10 coding. Both KUHR and NPR provided medical diagnosis data since 1 January 2010. DAR provided information on mortality.

### Definitions of severe COVID-19

**COVID-19 Hospitalization** was defined as hospital admission for at least 24 hours and i) a primary diagnosis with ICD-10 code U071, U072, or U109; or ii) a secondary diagnosis with ICD-10 code U071, U072, or U109 combined with a primary code of respiratory illness (defined as any of the ICD-10 J-codes). This definition was based on prior research in Norway [14].

**Death due to COVID-19** was defined as individuals with either i) COVID-19 listed as the primary cause of death (ICD-10 codes U071, U072, or U109 or ii) with a COVID-19 hospitalization in the 7 days preceding date of death. In the Dutch data registry, only the second definition was used, because cause of death was not available in death registry.

### Risk classifications for severe COVID-19

**The Dutch risk classification** was obtained from the ‘Adviesnota Vaststelling volwassen medische risicogroepen COVID-19-vaccinatiecampagne’ [‘Recommendation on establishing adult medical risk groups for the COVID-19 vaccination campaign’] in February 2023 [4].

This classification contains a list of chronic medical condition categories defined as either high- or moderate-risk for severe COVID-19. Each medical condition was independently coded using ICD-10 by two authors (AvB and IW), with no discrepancies found.

**The Norwegian risk classification** was obtained from the Norwegian Health Institute in July 2023 [5]. This scheme includes chronic medical condition categories with corresponding ICPC-2 and ICD-10 codes classified as either high-risk or increased-risk, which we renamed as high- and moderate-risk for consistency with other classifications.

**The European risk classification** was obtained from the ‘Core protocol for ECDC studies of COVID-19 vaccine effectiveness against hospitalization with Severe Acute Respiratory Infection (SARI), laboratory-confirmed with SARS-CoV-2 or with seasonal influenza – Version 3.0’ dated February 2024 [6]. This protocol provides ICD-10 codes for medical condition categories that are mandatory or optional for reporting in the ECDC studies. We classified the former medical condition categories as high-risk and the latter as moderate-risk.

Individuals in the Dutch or Norwegian datasets were assigned to have moderate- or high-risk chronic medical conditions if at least one relevant ICD-10 (both datasets) or ICPC-2 code (Norwegian cohort only) was recorded for that individual within the six months prior to 1 January 2020. For cancer diagnoses, records within five years prior to 1 January 2020 were used. These timeframes were defined according to the criteria specified in the classifications. Different medical risk classifications may use different codes for the same chronic medical condition, as each classification was developed with its own coding system. The ICD-10 and ICPC-2 codes used for this analysis are detailed in **Supplementary Tables 1-3**.

**Table 1.**
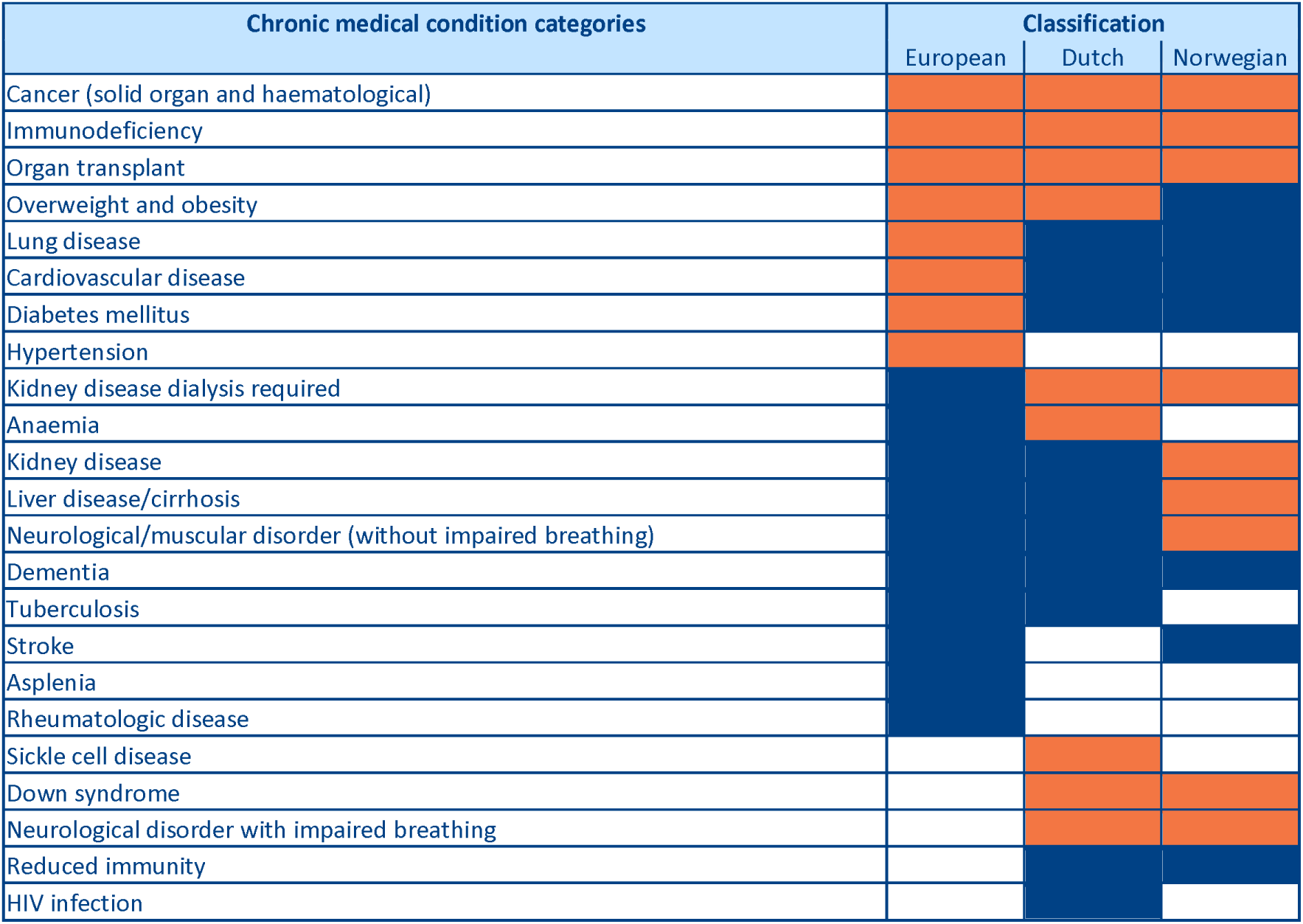
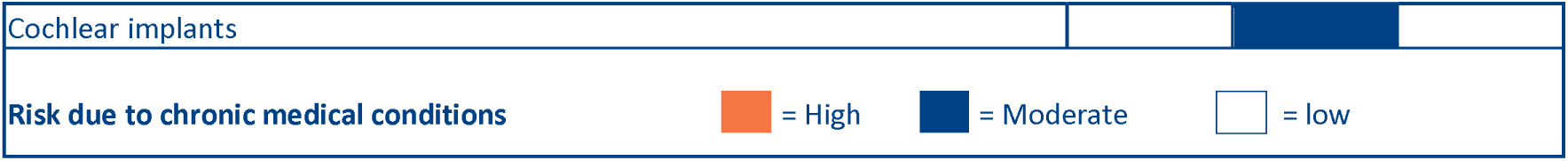
Comparison of European and national risk classifications for severe COVID-19. Chronic medical condition categories based on ICD-10 and ICPC-2 codes (see **Supplementary Tables 1-3**) and their respective risk groups for severe COVID-19 are shown. Different medical risk classifications may use different codes for the same chronic medical condition.

**Table 2:** Cumulative COVID-19 hospitalizations and deaths in 2020. Risk groups were defined on January 1, 2020. Individuals born after ary 1, 2020, are not included.

| Classification | The Netherlands |  |  |  |  | Norway |  |  |  |  |
| --- | --- | --- | --- | --- | --- | --- | --- | --- | --- | --- |
|  | General population | Hospitalization |  | Death |  | General population | Hospitalization |  | Death |  |
|  | n | n | Cumulative Incidence | n | Cumulative Incidence | n | n | Cumulative Incidence | n | Cumulative Incidence |
| National classification |  |  |  |  |  |  |  |  |  |  |
| - Low-risk | 15,171,220 | 22230 | 0.15% | 3004 | 0.02% | 4,708,944 | 1170 | 0.03% | 199 | 0.00% |
| - Moderate-risk | 1,973,422 | 13444 | 0.68% | 3285 | 0.17% | 706,399 | 786 | 0.11% | 225 | 0.03% |
| - High-risk | 257,637 | 3068 | 1.19% | 742 | 0.29% | 165,361 | 272 | 0.16% | 90 | 0.05% |
| European classification |  |  |  |  |  |  |  |  |  |  |
| - Low-risk | 15,671,292 | 23174 | 0.15% | 3069 | 0.02% | 4,995,283 | 1433 | 0.03% | 258 | 0.01% |
| - Moderate-risk | 165,325 | 865 | 0.52% | 208 | 0.13% | 72,122 | 71 | 0.10% | 24 | 0.03% |
| - High-risk | 1,565,662 | 14703 | 0.94% | 3754 | 0.24% | 513,299 | 724 | 0.14% | 232 | 0.05% |

**Table 3:** Performance COVID-19 classifications for severe COVID-19. Best performing risk-classification in bold.

|  | National classification |  |  |  | European classification |  |  |  | Brier score/MSE (SD) |  |  | AUC (95% CI) |  |  |
| --- | --- | --- | --- | --- | --- | --- | --- | --- | --- | --- | --- | --- | --- | --- |
|  | Sens | Spec | PPV | NPV | Sens | Spec | PPV | NPV | National class. | European class. | P-value | National class. | European class. | P-value |
| **The Netherlands** |  |  |  |  |  |  |  |  |  |  |  |  |  |  |
| Hospitalizations | **42.2%** | 87.2% | 0.70% | 99.9% | 39.7% | **90.1%** | **0.90%** | 99.9% | 0.002132 (0.04585) | **0.002131 (0.04582)** | 0.38 | 0.649 | **0.650** | 0.13 |
| Deaths | **57.3%** | 87.2% | 0.18% | >99.9% | 56.4% | **90.1%** | **0.23%** | >99.9% | 0.000404 (0.02005) | **0.000403 (0.02004)** | <0.001 | 0.725 | **0.733** | <0.001 |
| **Norway** |  |  |  |  |  |  |  |  |  |  |  |  |  |  |
| Hospitalizations | **45.6%** | 84.4% | 0.12% | >99.9% | 32.9% | **89.5%** | 0.12% | >99.9% | **0.00039395 (0.01982)** | 0.00039398 (0.01983) | <0.001 | **0.651** | 0.612 | <0.001 |
| Deaths | **63.2%** | 84.4% | 0.04% | >99.9% | 47.1% | **89.5%** | 0.04% | >99.9% | **0.0000920796 (0.00959)** | 0.0000920824 (0.00959) | <0.001 | **0.741** | 0.684 | <0.001 |

Abbreviations: Sens: Sensitivity, Spec: Specificity, PPV: Positive Predicted Value, NPV: Negative Predicted value, MSE: Mean Squared Error, AUC: Area Under the Curve

Individuals with one or more high-risk chronic medical condition categories, regardless of whether they also had one or more moderate-risk conditions, were defined as being at high-risk for severe COVID-19. Individuals with one or more moderate-risk chronic medical condition categories and no high-risk chronic medical condition were defined as being at moderate-risk for severe COVID-19. Individuals without any listed chronic medical condition categories were defined as low-risk. Pregnancy was not considered a criterion for categorizing individuals into the high- or moderate-risk groups in all classifications.

### Statistical Analysis

Data for the Netherlands and Norway were analyzed separately. Descriptive analyses were performed to compare the respective national with the ECDC classification. The prevalence of high- and moderate-risk medical condition categories, and the most frequent combinations of medical condition categories, were shown in upset plots which visualize co-occurance of chronic medical condition categories in the population. Due to privacy reasons, numbers below 10 for the Dutch data and below 5 for the Norwegian data were not shown.

We reported both absolute numbers and proportions of individuals by 10-year age groups, classified into high-, moderate-, and low-risk groups for severe COVID-19. The discordance between the European and each national classification was calculated as the proportion of the population with a different European versus national risk status for the full national population, COVID-19 hospitalized population, and COVID-19 deceased population, and visualized with matrix plots. Different medical risk classifications may use different codes for the same chronic medical condition because they were formulated as such by the classifications. The 95% confidence intervals (CI) of discordance and concordance proportions were calculated using the Clopper-Pearson method [15]. Moreover, cumulative incidence of COVID-19 hospitalizations and deaths within the full national populations were presented.

Logistic regression models were applied for each classification to predict hospitalization and death from COVID-19 in each country. Individuals were dichotomized into high- or moderate-risk versus low-risk groups, and classification performance was evaluated using sensitivity, specificity, positive predictive value (PPV), and negative predictive value (NPV). Additionally, model performance was compared with Brier scores (i.e., mean squared errors) of the national and European classifications. Differences between these scores were tested using the Wilcoxon signed-rank test. Moreover, the Area Under the Curve (AUC) values were computed and statistically compared using DeLong’s test to determine if there were significant differences.

To understand the factors related to discordance, we visualized the co-occurance of chronic medical condition categories in the discordant general, hospitalized, and deceased populations with upset plots. Also, to understand how coding differences affect overall discordance, we reviewed the most prevalent diseases in hospitalized and deceased populations one by one by assessing the differences in population sizes between classifications. Since the Netherlands and Norway consider individuals above a certain age as high-risk for severe COVID-19, we also assessed the discordance in the general population in three scenarios: 1) the entire population regardless of age, 2) individuals younger than 60 years of age, aligning with the Dutch schemes [16], and 3) individuals younger than 65 years of age, aligning with the Norwegian schemes [5].

A sensitivity analysis was conducted to assess the importance of the reference date by evaluating the discordance in the general population for 1 January 2018, 1 January 2019, and 1 January 2021, instead of 1 January 2020.

### Code Availability

The codes reproducing the results of this study are publicly available at https://github.com/IWesterhof/COVID-19-risk-classifications.

### Data Availability

The aggregated tables of counts generated and analyzed in this study are publicly available at https://github.com/IWesterhof/COVID-19-risk-classifications.

Results are based on calculations by the authors using non-public microdata from Statistics Netherlands. Under certain conditions, these microdata are accessible for statistical and scientific research. For further information.

### Ethics

The analyses conducted with data from the Netherlands did not fall under the scope of the Dutch Medical Research Involving Human Subjects Act (WMO) and, therefore, did not require approval from an accredited ethics committee in the Netherlands. For the analyses conducted with data from Norway, the Regional Committee for Medical and Health Ethics of South/East Norway (no. 285687) approved the study. The Norwegian Data Protection Services for research and the University of Oslo approved the Data Protection Impact Assessment – DPIA (no. 341884).

## Results

The Dutch cohort contained records of 17.4 million individuals (median age 42, interquartile range (IQR) 22-60), while the Norwegian cohort contained records of 5.6 million individuals (median age 39, IQR 21-57).

### Comparison of risk classifications for severe COVID-19

Chronic medical condition categories included in the European, Dutch, and Norwegian risk classifications and their respective risk groups for severe COVID-19 are shown in **Table 1**.

Several chronic medical condition categories were consistently considered high-risk (e.g., cancer, immunodeficiency, and organ transplants) and moderate-risk (e.g., dementia) in the European and national classifications. There was some variability in the risk classifications of other conditions. Obesity was considered high-risk in the European classification but moderate-risk in both national classifications; lung disease, cardiovascular disease, and diabetes mellitus were considered high-risk in the European classification but moderate-risk in both national classifications. Hypertension was only listed in the European classification. Down syndrome and neurological disorders with impaired breathing were not included in the European classification but were considered high-risk in both national classifications.

### Classification of the general population by risk for severe COVID-19

Based on the national classifications, the most prevalent chronic medical condition categories in the Netherlands were lung disease, followed by cardiovascular disease, reduced immunity, and neurological/-muscular disorder; all classified as moderate-risk (**Figure 1 A**). In Norway, the most prevalent conditions were cardiovascular disease, lung disease, and diabetes mellitus; these were also classified as moderate-risk (**Figure 1 B**). Based on the European classification, the most prevalent chronic medical condition categories in the Netherlands were cardiovascular disease, hypertension, and cancer, and in Norway cardiovascular disease, diabetes mellitus, and cancer; all classified as high-risk (**Figure 1 C-D**). Differences in the prevalence between classifications arise from variations in the diagnosis codes to define a specific medical risk condition. For example, in the Netherlands, 0.54% of the population was classified as having cancer using the national classification, compared to 2.0% using the European classification because this classification included seven additional ICD-10 codes (**Supplementary Tables 1-2**). In Norway, 2.3% of the population was classified as having cancer using the national classification compared to 2.1% using the European classification due to differences in included codes.

**Figure 1.**
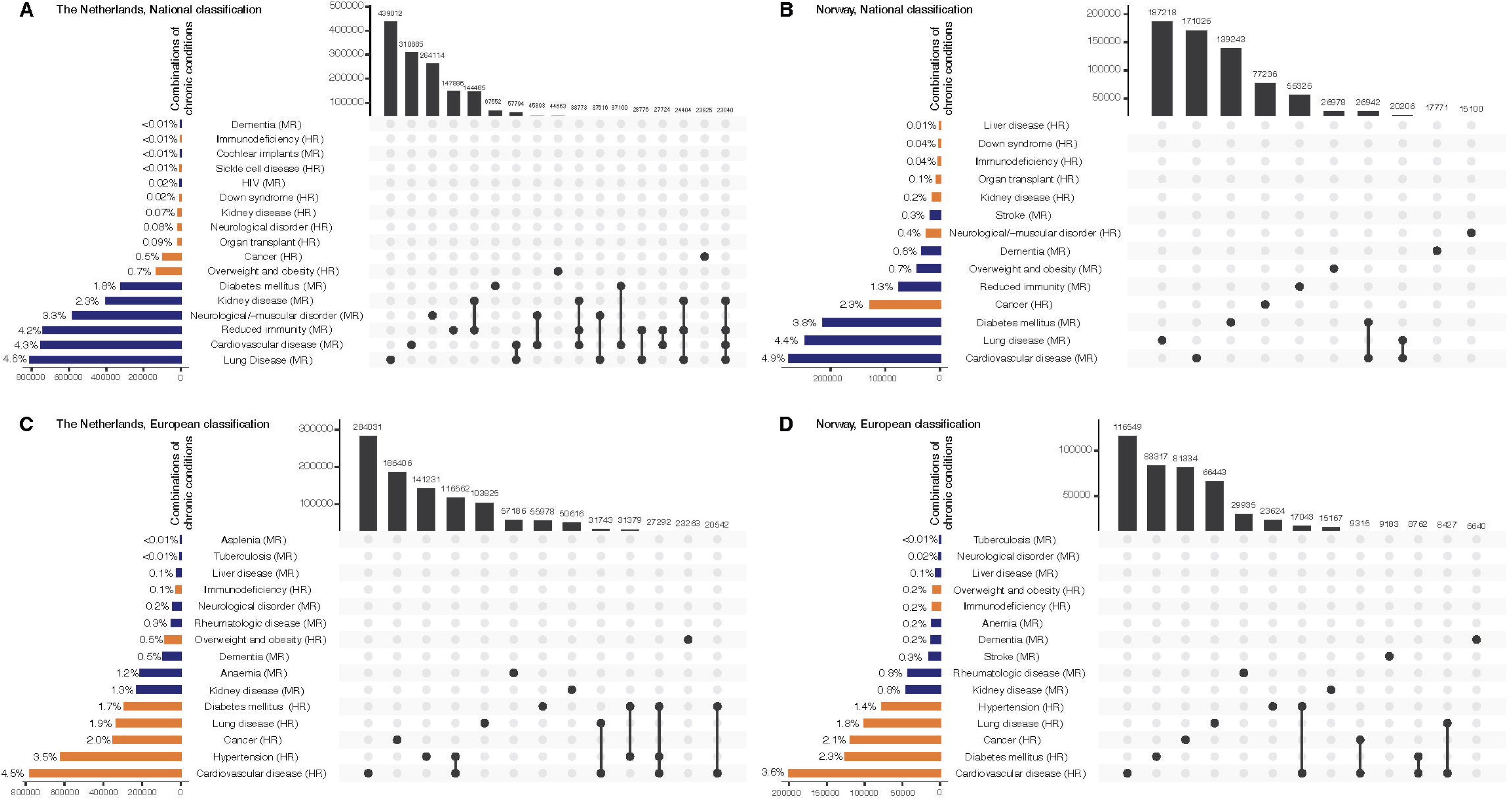
Chronic conditions in the general population. **A-B)** National and **C-D)** European medical risk classification for severe COVID-19. In all classifications, chronic medical condition categories were assigned if registered in the past six months prior to 1 January 2020, except for cancer which was assigned if registered in the prior five years. The left bars show the number and proportion of each chronic medical condition category in the population. Orange bars indicate the high-risk chronic conditions according to the classification, while blue bars indicate moderate-risk chronic conditions. The dots represent the presence of one or more chronic medical condition categories in an individual, with the upper bars showing the combinations that occur in at least 1% of the general population.

In both countries and across all classifications, most individuals under 80 years of age were assigned low-risk for developing severe COVID-19 (**Figure 2**). Among children and adolescents (under 20 years old), 92.8% were classified as low-risk, 6.9% as moderate-risk, and 0.3% as high-risk, according to national schemes, while this was 98.5%, 0.4%, and 1.2%, respectively, according to the European classification. In Norway, the national classification identified 98.5% as low-risk, 0.4% as moderate-risk, and 1.2% as high-risk, while this was 96.4%, 0.5%, and 3.1%, respectively, according to the European classification. In both countries and across all classifications, the percentage of individuals in moderate- and high-risk groups increased with age, reaching 41.1% and 53.6% for those over 80 years in the national classifications, and 41.8% and 37.7% in the European classifications in the Netherlands and Norway, respectively.

**Figure 2.**
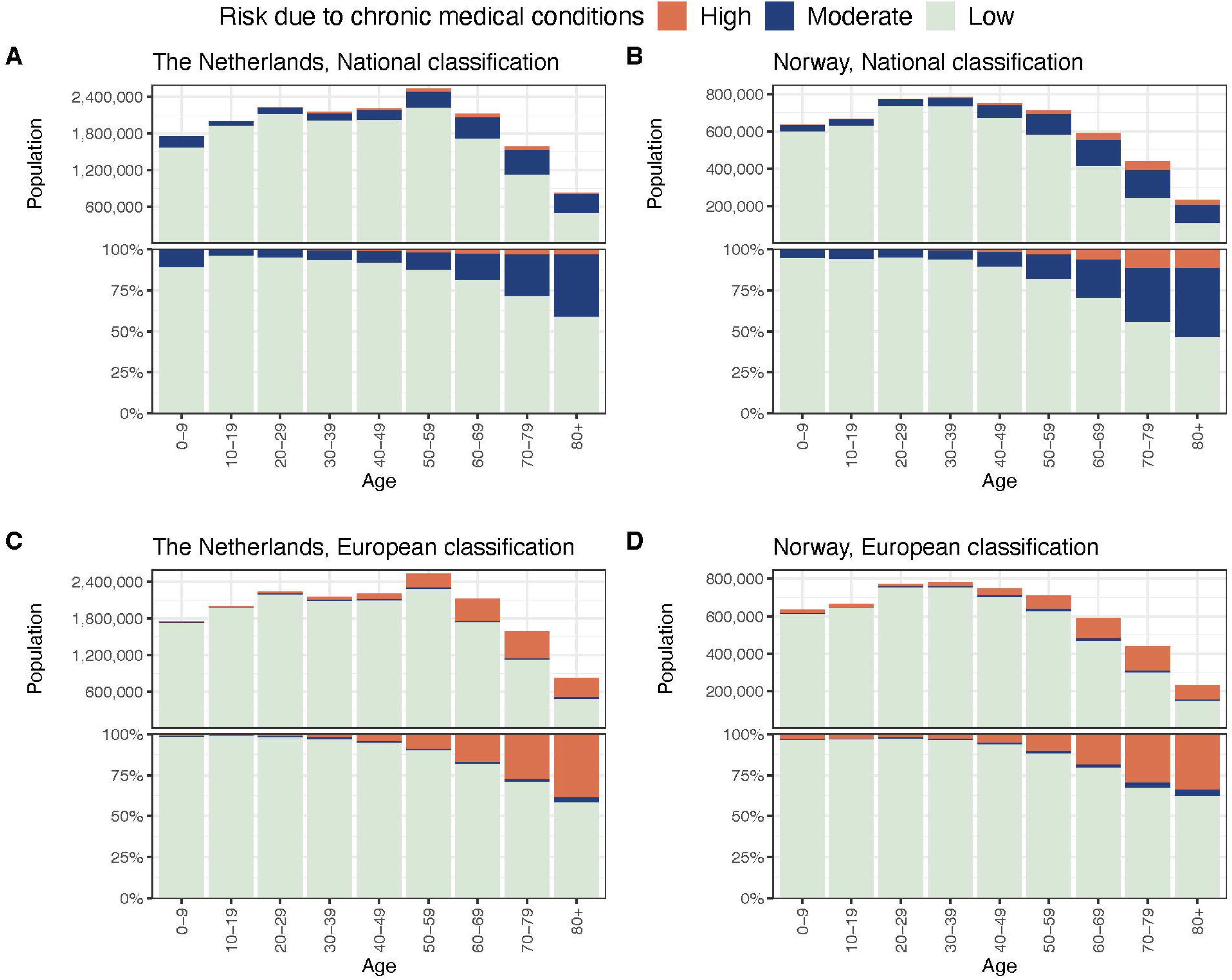
Classification of the general population by risk for severe COVID-19. Population shown as absolute numbers (top) and proportions (bottom) classified by age and risk for severe COVID-19 due to chronic medical condition categories in the **A-B)** national and **C-D)** European classifications.

In the Netherlands and Norway, risk classification differed for 12.0% and 13.8% of the general population, respectively, between the national and the European classification schemes (**Figure 3 A-B**). The European classification assigned more people as high-risk (9% in the Netherlands, 9.2% in Norway) compared to national classifications (1.5% and 3.0%, respectively). Conversely, the national classifications identified more people as moderate-risk (11.3% and 12.7%, respectively) than the European classification (1.0% and 1.3%, respectively). According to the national classifications, 87.2% of the population in the Netherlands and 84.4% in Norway were classified as low-risk. These percentages were slightly higher using the European classification (90.1% and 89.5%, respectively).

**Figure 3.**
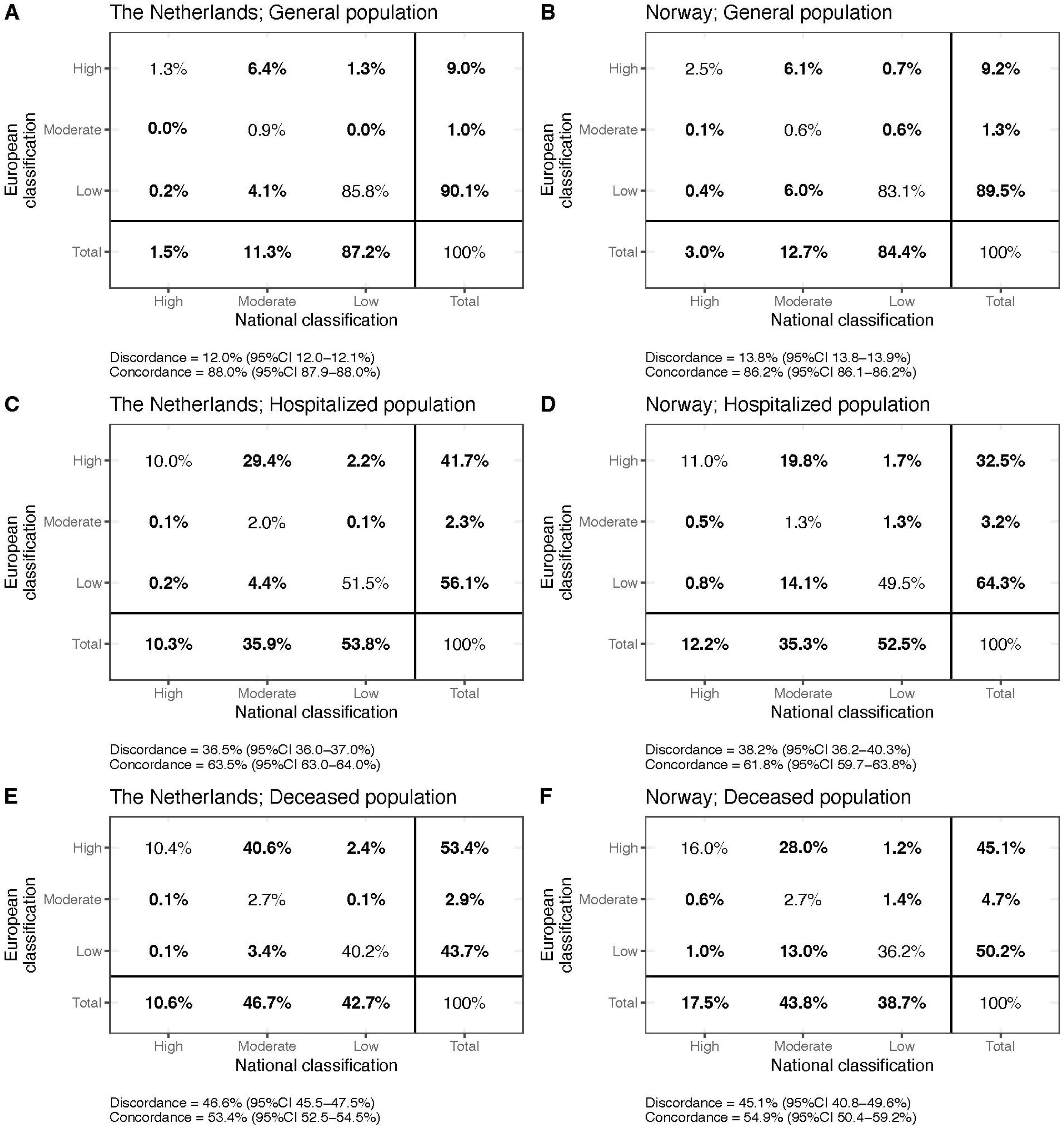
Discordance in assigning risk status to individuals between the European and national classifications in the A-B) general population, **C-D)** hospitalized population, and **E-F)** deceased population. The discordance between the European and each national classification was calculated as the proportion of the general populations with a different risk status (off-diagonal elements). Concordance is the proportion of the population classified into the same risk group (diagonal elements). Since every individual belongs to one of the risk groups, the elements of the matrix plots add up to 100%.

### Discordance for population with severe COVID-19 in 2020

The Netherlands had 39,063 hospitalizations (median age 70 years [IQR 58-78]) and 7,031 deaths (median age 78 years [IQR 71-83]). In Norway, there were 2,228 hospitalizations (median age 64 years [IQR: 51-78]) and 514 deaths (median age 84 years [IQR: 75-90]).

The cumulative incidence of COVID-19 hospitalizations and deaths increased with higher risk levels for both the national and European classifications (**Table 2**). For example, the cumulative incidence of hospitalizations was 0.15% for the low-risk group, 0.68% for the moderate-risk group, and 0.14% for the high-risk group for the national classification compared to 0.15%, 0.52%, and 0.94%, respectively, for the European classification.

Similarly, the cumulative incidence of COVID-19 deaths was 0.02% for low-risk, 0.17% for moderate-risk, and 0.29% for high-risk groups for the national classification compared to 0.02%, 0.13%, and 0.24%, respectively, for the European classification.

Cardiovascular disease, classified as high-risk in the European classification and moderate-risk in the national classifications, is the most prevalent chronic condition among hospitalized and deceased individuals (**Supplementary Figure 1-2**). Another important disease that is prevalent and differently classified is Diabetes Mellitus.

For all ages and both countries, and similar to the general population, the European classification assigns more individuals to high-risk, while national classifications define more individuals as moderate-risk (**Supplementary Figures 3-4**). Discordance was even higher among hospitalized (the Netherlands 36.5% and Norway 38.2%), and deceased (the Netherlands 46.6% and Norway 45.1%) populations, compared to the general population (discordance in the Netherlands 12.0% and Norway 13.8%), as shown in **Figure 3C-F**. Moreover, a large part of the hospitalized (51.5% in the Netherlands and 49.5% in Norway) and deceased (40.2% and 36.2%, respectively) populations was defined as low-risk in both classifications, which means these individuals had none of the chronic medical conditions listed in classifications.

National classifications showed consistently higher sensitivity than European classifications (**Table 3**). For example, in Norway, sensitivity for hospitalization was 45.6% for the national classification versus 32.9% for the European classification; for COVID-19 deaths, sensitivity was 63.2% versus 47.1%, respectively. Specificity was always higher for the European classification. PPV and NPV were comparable across classifications, with very low PPVs (≤0.9%) and NPVs near 100%. The Brier scores for all classifications were below 0.01, consistent with the low incidence of outcomes and the correspondingly low predicted risks. Discrimination and calibration metrics (Brier score and AUC) favored the national classification in Norway (both p<0.001), while in the Netherlands, no significant differences were seen for hospitalizations (p≥0.13), and the European classification performed better for COVID-19 mortality (p<0.001).

Comparison between the Norwegian and Dutch classifications for the hospitalized and deceased populations in Norway shows a discordance of 17.1% and 21.8%, respectively (**Supplementary Figure 5**). This is mainly caused by differences in selecting diseases (ICD-19 and ICPC-2 codes) within the disease categories lung disease, cardiovascular disease, and dementia.

### Discordance between risk classifications and the related factors

Discordance was higher in elderly because the number of individuals with one or more medical risk conditions was higher than in younger age groups (**Figure 3-4; Supplementary Figure 6**). **Figure 4** shows the chronic medical condition categories in the general population who were classified differently by the European and respective national classification schemes (the discordant population). In the discordant population in the Netherlands, 23.5% had neurological/-muscular disorders, 23.8% reduced immunity, and 13.6% kidney disease according to the national classification (**Figure 4A**). In the discordant population in Norway, 6.7% had reduced immunity, and 2.3% neurological/muscular disorders, classified as moderate- and high-risk, respectively (**Figure 4B**). On the contrary, the European classification included chronic medical condition categories that were not included in the national classifications. In the Netherlands, examples were hypertension and kidney disease with prevalences of 25.5% and 6.3% in the discordant population. In Norway, 8.2% of the discordant population had hypertension and 3.6% had kidney disease according to European schemes, but not included in national schemes. Unlike Norway, none of the discordant individuals in the Netherlands were classified as overweight/obese or diabetic based on the European classification. Similar was observed for the hospitalized and deceased discordant populations (**Supplementary Figures 7-8**).

**Figure 4.**
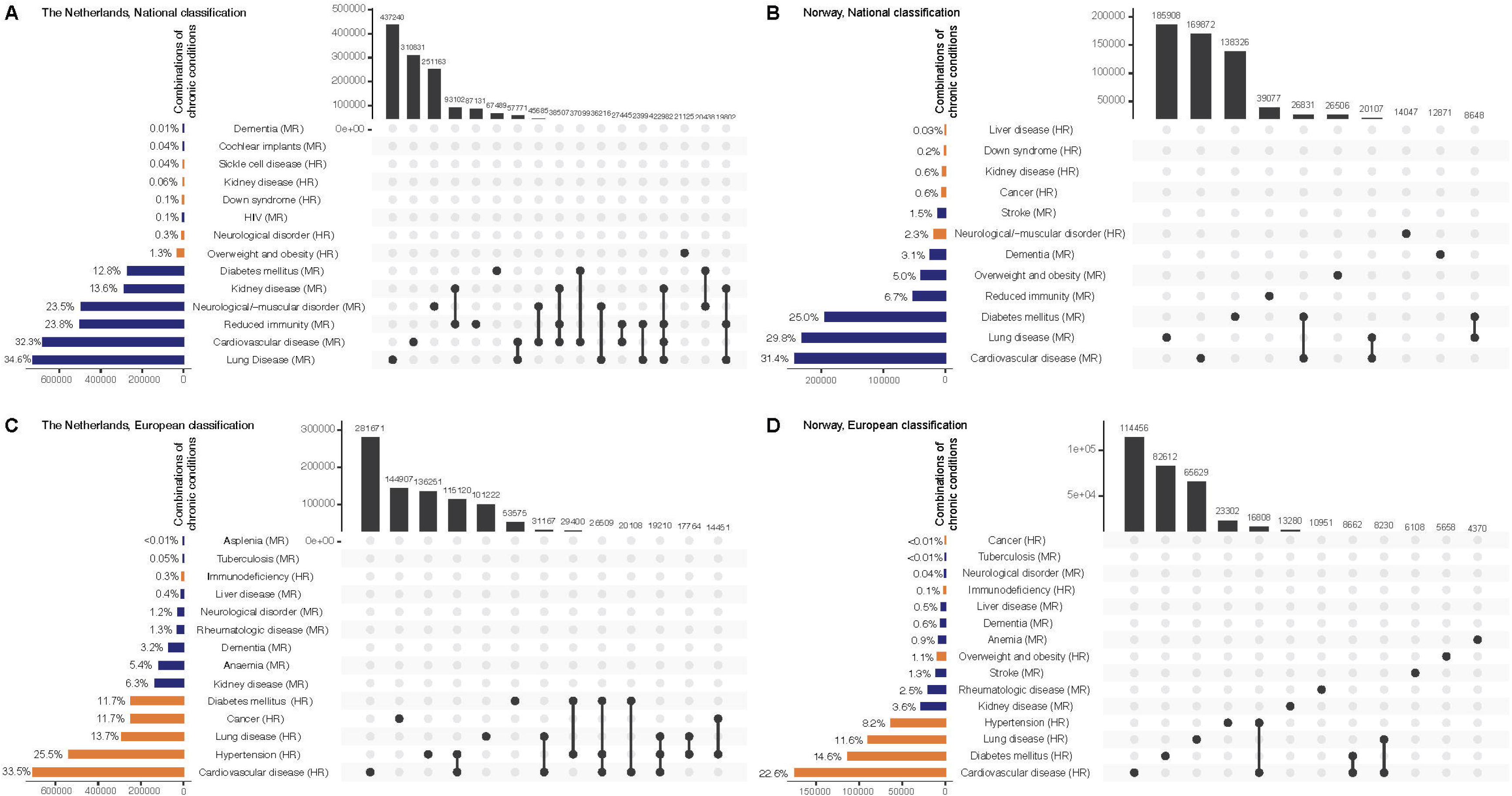
Chronic conditions in the discordant general population. A-B) National and **C-D)** European medical risk classification for severe COVID-19. In all classifications, chronic medical condition categories were those that had been registered in the past six months prior to 1 January 2020, except for cancer which was included as a chronic medical condition if registered in the past five years preceding 1 January, 2020. The dots represent the presence of one or more chronic medical condition categories. The left bars show the number and proportion of each chronic medical condition category in the population. Orange bars indicate the high-risk chronic conditions according to the classification, while blue bars indicate moderate-risk chronic conditions. The upper bars show the combinations of chronic conditions that occur in at least 1% of the discordant general population. [6][4][5]

Some of the discordance in the most prevalent chronic medical condition categories (lung disease, cardiovascular disease, diabetes mellitus, cancer, and kidney disease) among hospitalized and deceased populations may stem from differences in ICD-10 and ICPC-2 codes. In the Netherlands, the national classification identified more individuals with lung disease and diabetes, while the Norwegian system captured more cases across all disease categories except kidney disease, indicating that it includes more ICD-10 codes (**Supplementary Table 5**). The European classification identified more individuals with cardiovascular disease and cancer in the Netherlands and more individuals with kidney disease in Norway. Notably, in the Netherlands, cancer was better captured with the European classification, identifying more cases in the general (2.0% versus 0.5%), hospitalized (9.1% versus 4.1%), and deceased (10.8% versus 4.1%) populations compared to the Dutch classification. Similarly, in Norway, the European classification identified more individuals with kidney disease in the general (0.8% versus 0.2%), hospitalized (10.1% versus 2.4%), and deceased (13.3% versus 2.3%) populations compared to the national classification.

In the Netherlands, the most common chronic medical condition categories varied by age group (**Supplementary Figures 9-10**). In those younger than 60 or 65, lung disease was the most prevalent condition, followed by neurological/-muscular disorder and cardiovascular disease. In Norway, the national classification showed no differences between all ages and individuals younger than 65 and 60 years of age in most pronounced chronic conditions. In the Netherlands, based on the European classification there was no change in the order of most prominent conditions contributing to discordance in the population aged below 60 years compared to all ages. In contrast, for those aged below 65 years, the most prevalent conditions were cardiovascular disease, diabetes mellitus, and lung disease, but not hypertension and cancer.

#### Sensitivity analyses

No meaningful differences in discordance percentages were observed when using different reference dates to define the study populations (data not shown).

## Discussion

In this multi-country, nationwide registry-linkage study, we compared the respective national with the European medical risk classification for severe COVID-19, and classified the populations of the Netherlands and Norway according to this risk, across different age groups. For clarity, we reiterate that individuals with at least one high-risk chronic condition were classified as high-risk, those with only moderate-risk conditions as moderate-risk, and those without listed conditions as low-risk (see Methods for details). Our findings revealed substantial differences between the national and European risk classifications in the general population (discordance in the Netherlands 12.0% and Norway 13.8%), hospitalized patients (the Netherlands 36.5% and Norway 38.2%), and deceased patients (the Netherlands 46.6% and Norway 45.1%), with the European classification scheme assigning a broader range of chronic medical condition categories to the high-risk category than the two national classifications. Consequently, a larger proportion of the population is classified as high-risk when using the European classification compared to the national classifications. Conversely, a larger proportion of the population is classified as moderate-risk when using the respective national classifications compared to the European classification. The Dutch national classification may be underinclusive for cancer and cardiovascular disease, and the Norwegian national classification for kidney disease, while those disease categories are probably more clinically relevant as they are prevalent among hospitalized and deceased populations. Interestingly, a large part of the hospitalized (51.5% in the Netherlands and 49.5% in Norway) and deceased (40.2% and 36.2%) populations were defined as low-risk in both national and European classifications. This may be due to chronic medical conditions not captured by either classification. This suggests that the classifications differ even more when applied to severe COVID-19 outcomes and fail to identify a considerable share of affected individuals. Moreover, our results indicate that the national classifications provide better sensitivity, while the European classifications offer better specificity. Additionally, the accuracy and discrimination of individuals at risk for severe COVID-19 outcomes is greater for the national classification in Norway; however, this was not demonstrated for the national classification in the Netherlands. Notably, none of the classifications showed significantly better accuracy or discrimination for predicting hospitalizations. In contrast, the European classification was significantly better in identifying individuals at risk of death due to COVID-19.

These discrepancies may have implications for COVID-19 vaccination programs, which prioritize specific risk populations. Although primarily designed for research focusing on COVID-19 vaccine effectiveness and enhancing SARI surveillance in Europe, the European protocols have influenced public health policy in the past, as demonstrated, for example, with influenza vaccination strategies [17]. However, we recognize that this classification is primarily surveillance-oriented and not an officially mandated European clinical standard.

While not a universal policy tool, this classification provides a structured framework that can support public health decision-making alongside other clinical and epidemiological considerations. Therefore, the European classification for severe COVID-19 could remain relevant for broader public health decisions and policy. Our comparative analysis suggests that using the European classification, 9.0% of the population in the Netherlands and 9.2% of the population in Norway are high-risk due to chronic medical condition categories and should be vaccinated, while these percentages are only 1.5% and 3.0%, respectively, when using the national classifications. Vaccination programs that prioritize both moderate- and high-risk groups, the European classification would result in 10.0% of the Dutch population and 10.5% of the Norwegian population having to be vaccinated, compared to 12.8% and 13.7%, respecitively, if using the respective national classifications. Moreover, chronic medical condition categories are defined based on different ICD-10 and ICPC-2 codes in the three classifications, resulting in differences in prevalence of individual medical risk categories across classifications. Therefore, the choice between the European or national classifications may have important consequences for vaccination prioritization and coverage, potentially influencing the impact of vaccination campaigns in preventing severe COVID-19.

In our analyses, the national classification performed better in Norway for both COVID-19 hospitalization and mortality, while in the Netherlands, the European classification performed better for mortality, with no clear differences observed for hospitalization. Although the statistical performance measures provide useful comparison, they highlight that no single classification is universally optimal. Sensitivity was consistently higher for national classifications, indicating better identification of individuals at high- or moderate-risk of severe COVID-19 outcomes. In contrast, the European classification demonstrated higher specificity, more accurately excluding low-risk individuals who did not experience severe outcomes. Given the potential severity of COVID-19 and the high effectiveness of vaccines alongside the rarity of serious adverse events [18], it may be preferable to prioritize sensitivity when designing vaccination policies to ensure that individuals with chronic medical conditions are not overlooked.

The dominant medical condition categories generating the discordance in risk status were lung and cardiovascular disease in both Norway and the Netherlands, neurological disorders in the Netherlands, and diabetes in Norway. Hypertension was one of the most pronounced discordant chronic condition categories between classifications. According to the European classification, but not to the national ones, hypertension was considered a high-risk disorder for severe COVID-19, and its prevalence estimate was 3.5% in the Netherlands and 1.4% in Norway. Several studies have indicated that hypertension was not an independent predictor of severe COVID-19 [19,20]. Therefore, the high-risk population might be overestimated in the European classification. Lung disease, cardiovascular disease, and diabetes mellitus were also among the most pronounced medical condition categories leading to discordance, which were diagnosed in 1.7-4.6% of the population according to the national and European classifications. These conditions were classified as moderate-risk in the national classifications but as high-risk in the European classification. Although multiple studies have shown an increased risk of severe COVID-19 in individuals with these medical condition categories [21–23], it is unknown whether they should be part of the high- or moderate-risk group. Interestingly, the European classification does not include Down syndrome and neurological/-muscular disorder as risk factors for severe COVID-19, while the national classifications consider them as moderate- or high-risk chronic conditions. Several studies have demonstrated an increased risk of COVID-19 hospitalization and death among individuals with Down syndrome [24,25] and multiple sclerosis [26,27]. However, our findings show that only a very small fraction of the discordant population has Down syndrome (≤0.2% in both countries), while those with neurological/muscular disorders make up a larger proportion—23.5% in the Netherlands and 2.3% in Norway. This supports the need to assess the discordant conditions in relation to severe COVID-19 to ensure vaccination prioritization and coverage of individuals at high-risk.

Our study has several limitations. First, the available data did not allow us to assess how well the classifications predict severe COVID-19 outcomes after 2020, when vaccination became available and reinfections were frequently observed. As individuals were prioritized for vaccination based on the national guidelines which align with the data used in our study, this would introduce bias. Second, due to limited testing capacity in 2020, we were unable to clearly identify exposed individuals and analyze their risk of severe COVID-19 outcomes upon infection. Therefore, we assessed the risk of severe COVID-19 for the entire national population rather among those infected. Lastly, a limitation specific to the Norwegian data is the absence of ICD-10 codes for asplenia in the registries. However, since asplenia is a very rare condition, estimated at 0.44 cases of surgical asplenia per 100,000 person-years in Norway, this limination is unlikely to affect our overall conclusions [28].

Future research should aim to establish a consensus on which chronic medical condition categories are universally recognized as conferring moderate- and high-risk for severe COVID-19, especially in the post-pandemic era, where the population has substantial immunity due to vaccination and infections. We propose a matched multi-country cohort study to estimate the risk of hospitalization and death due to COVID-19 in individuals with and without specific chronic conditions, accounting for confounders like age, vaccination status, and circulating variants. Additionally, the importance of chronic medical conditions in the hospitalized and deceased populations that are not included in the national and European classifications, and therefore classified as low-risk, should be further assessed. This will help understand which additional conditions should be included in a risk classification for severe COVID-19. The analyses should ideally be conducted at the diagnostic code level, as our findings highlight substantial differences between classifications for specific disease categories. This approach will also help determine whether certain ICD-10 and ICPC-2 codes are overly broad or clinically unsubstantiated. This research will be crucial for deciding which classification most accurately captures the population at highest risk. Moreover, mathematical modeling studies can help to understand the impact of different risk classifications and optimal vaccination strategies [29–32]. Incorporating risk stratification into modeling studies has the potential to design more effective strategies, a consideration often overlooked in current research efforts. Furthermore, exploring emerging data sources and digital health technologies could improve the identification and monitoring of moderate- and high-risk populations.

## Conclusion

There are substantial differences between the European and the respective national medical risk classifications for severe COVID-19 based on pre-existing chronic medical condition categories, which may have important implications for vaccination strategies. The European classification suggests that a larger proportion of the population is at high-risk for severe COVID-19 and may require vaccination. Classification performance showed that no single classification is universally optimal. Further research should identify which chronic conditions are associated with risk of developing severe COVID-19 and should be included in risk classifications.

## Author Contributions

Conceptualization: I Westerhof, G Rozhnova. Data management: I Westerhof, A de Boer. Methodology: I Westerhof, A de Boer, G Rozhnova. Input statistical methodology: N Zuithoff. Investigation: I Westerhof, A de Boer. Formal analysis: I Westerhof, A de Boer. Visualization: I Westerhof. Writing—original draft: I Westerhof, A de Boer, G Rozhnova. Writing—review and editing: I Westerhof, A de Boer, A Lupattelli, I Slurink, N Zuithoff, O Boldea, HME Nordeng, JR Bosdriesz, F Pijpers, M Schim van der Loeff, M Knol, JHHM van de Wijgert, P Bruijning, and G Rozhnova.

## Acknowledgments

We are grateful to all the participating families in the Netherlands and Norway who are part of the health registries and made this research possible. GR, IW, AdB, and OB were supported by the ZonMw project (10430362220002). GR was supported by Fundação para a Ciência e a Tecnologia project 2022.01448.PTDC, DOI 10.54499/2022.01448.PTDC. GR, IW, AL, and PB were supported by the VERDI project (101045989), funded by the European Union. Views and opinions expressed are however those of the author(s) only and do not necessarily reflect those of the European Union or the Health and Digital Executive Agency. Neither the European Union nor the granting authority can be held responsible for them. IW was supported by the scholarship program iAPOGEE “International Alliance for PharmacoGenetic Epidemiology Excellence” at the University of Oslo, Norway. Data acquisition of the Norwegian data in this study was funded by HN’s EU-COVID-19 project, funded by the Norwegian Research Council’s COVID-19 Emergency Call (project no. 31270). The work on the Norwegian data was performed on the TSD (Tjeneste for Sensitive Data) facilities, owned by the University of Oslo, operated and developed by the TSD service group at the University of Oslo, IT-Department (USIT).

## Declaration of generative AI in scientific writing

During the preparation of this work the author(s) used Chat GPT in order to improve readability and language. After using this tool/service, the author(s) reviewed and edited the content as needed and take(s) full responsibility for the content of the publication.

