## Supplementary material for "Medical Risk Classification For Severe COVID-19 Based On Chronic Medical Conditions: A Comparative Analysis": Supplementary Figures Titles.docx

**Supplementary Figure 1. Chronic conditions in the hospitalized population. A-B)** National and **C-D)** European medical risk classification for severe COVID-19. In all classifications, chronic medical condition categories were those that had been registered in the past six months prior to hospitalization, except for cancer which was included as a chronic medical condition if registered in the past five years preceding the hospitalization. The dots represent the presence of one or more chronic medical condition categories. The left bars show the number and proportion of each chronic medical condition category in the population. Orange bars indicate the high-risk chronic conditions according to the classification, while blue bars indicate moderate-risk chronic conditions. The upper bars show the combinations of chronic conditions that occur in at least 1% of the hospitalized population.

**Supplementary Figure 2. Chronic conditions in the deceased population. A-B)** National and **C-D)** European medical risk classification for severe COVID-19. In all classifications, chronic medical condition categories were those that had been registered in the past six months prior to 1 January 2020, except for cancer which was included as a chronic medical condition if registered in the past five years preceding 1 January 2020. The dots represent the presence of one or more chronic medical condition categories. The left bars show the number and proportion of each chronic medical condition categories in the population. Orange bars indicate the high-risk chronic conditions according to the classification, while blue bars indicate moderate-risk chronic conditions. The upper bars show the combinations of chronic conditions that occur in at least 1% of the deceased population.

**Supplementary Figure 3. Classification of the hospitalized population by risk for severe COVID-19.** Population shown as absolute numbers (top) and proportions (bottom) classified by age and risk for severe COVID-19 due to chronic medical condition categories in the **A-B)** national and **C-D)** European classifications. Panels B and D do not show age group 0-9 due to low numbers. Grey bars indicate moderate and high risk together due to low numbers.

**Supplementary Figure 4. Classification of the deceased population by risk for severe COVID-19.** Population shown as absolute numbers (top) and proportions (bottom) classified by age and risk for severe COVID-19 due to chronic medical condition categories in the **A-B)** national and **C-D)** European classifications. Panels do not show age groups 0-39 due to low numbers. Grey bars indicate moderate and high risk together due to low numbers.

**Supplementary Figure 5. Discordance in assigning risk status to A) hospitalized and B) deceased individuals in Norway between the Norwegian and Dutch classifications.** The discordance between the Norwegian and Dutch classifications was calculated as the proportion of the general populations with a different risk status (off-diagonal elements). Concordance is the proportion of the population classified into the same risk group (diagonal elements). Since every individual belongs to one of the risk groups, the elements of the matrix plots add up to 100%.

**Supplementary Figure 6. Discordance in assigning risk status to individuals between the European and national classifications.** General population of **A-B)** ages below 65 years, and **C-D)** ages below 60 years. The discordance between the European and each national classification was calculated as the proportion of the general populations with a different risk status (off-diagonal elements). Concordance is the proportion of the population classified into the same risk group (diagonal elements). Since every individual belongs to one of the risk groups, the elements of the matrix plots add up to 100%.
