## Supplementary material for "Medical Risk Classification For Severe COVID-19 Based On Chronic Medical Conditions: A Comparative Analysis": Supplementary Table 1 .docx

**Supplementary Table 1** European risk classification, ICD-10 codes [6].

| **Risk group** | **Disease category** | **ICD-10 codes** |
| --- | --- | --- |
| High | Cardiovascular disease | A52.01, B37.6, B58.81, I05-09, I11, I13, I20-25, I26.9, I27, I30-51, I97.0-97.1, R00.1, T81.718A, T81.72XA, T82.817A, T82.818A, Q20-24, Q25.1-25.2, Q26.0-26.1, Q26.8, Q87.4, R01.1, R02 |
| High | Cancer | C00-96 |
| High | Diabetes mellitus | E10-11 |
| High | Hypertension | I10, I15, I15.1, I15.2, I15.8, I27.0, I97.3 |
| High | Immunodeficiency or organ transplant | B20, D80-84, D89.8-89.9, Z21, Z94 |
| High | Lung disease | A15, J18.2, J40-47, J60-94, J96, J99, M34.81, M05.10 |
| High | Obesity | E66.01, E66.2, E66.9 |
| Moderate | Anemia | D50-64 |
| Moderate | Asplenia *(not available in Norwegian registries)* | Q89.01, Q20.6, Z90.81 |
| Moderate | Dementia | F01, F03, F05, G30-31, G91, G94 |
| Moderate | Kidney disease | M10.30, N00-19, N20.0, N28.9 |
| Moderate | Liver disease | K70, K72-74, K75.4, K76.9 |
| Moderate | Neurological disorder | G70, G73.7 |
| Moderate | Rheumatologic diseases | M30-34, M35.5, M35.8-35.9, M05-06, M08, M12.00 |
| Moderate | Stroke | G93, I67.83, I69 |
| Moderate | Tuberculosis | A15-19 |
