## Supplementary material for "Medical Risk Classification For Severe COVID-19 Based On Chronic Medical Conditions: A Comparative Analysis": Supplementary Table 2.docx

**Supplementary Table 2** Dutch risk classification, ICD-10 codes [4].

| **Risk group** | **Disease category** | **ICD-10 codes** |
| --- | --- | --- |
| High | Cancer | C00-C75, C81-86, C88, C90-96. |
| High | Down syndrome | Q90 |
| High | Immunodeficiency disease | D800.0, D81, D830.0-830.2, D83.8-83.9 |
| High | Kidney disease (dialysis required) | N18.5 |
| High | Neurological disorders compromising respiration | G00-G99 in combination with R06 |
| High | Organ transplantation | Z94.0-94.4, Z94.8 |
| High | Overweight and obesity | E66 |
| High | Sickle cell disease | D57.0-57.2 |
| Moderate | Cardiovascular disease | C38, C49.3, D511, D213, I00-2, I05-9, I20-3, I30-52, Q20-8, P29.3, Z950 |
| Moderate | Chronic kidney disease | N00-N39 |
| Moderate | Cochlear implants | Z96.2 |
| Moderate | Dementia | F00-01, F03, F10.6 |
| Moderate | Diabetes mellitus | E10-14 |
| Moderate | HIV | B20-24 |
| Moderate | Lung disease | A15-19, C30-34, C39, D86, E84, J00-99, Q30-34 |
| Moderate | Neurological/-muscular disorders | G00-99, I60-69 |
| Moderate | Reduced immunity | B17.0, B18.0-18.1, C81-96, D50-89, E24-27, K50-51, K70-77, M0-14, M353, N0-39. |
