## Supplementary material for "Medical Risk Classification For Severe COVID-19 Based On Chronic Medical Conditions: A Comparative Analysis": Supplementary Table 3.docx

**Supplementary Table 3** Norwegian risk classification, ICD-10 and ICPC-2 codes[5].

| **Risk group** | **Disease category** | **ICD-10 codes** | **ICPC-2 codes** |
| --- | --- | --- | --- |
| High | Cancer | C0-7, C80-96, D45, D47 | - |
| High | Down Syndrome | Q90 | - |
| High | Immunodefiency disease | D80-84 | - |
| High | Organ transplantation | Z94.0-94.4, Z94.8 | - |
| High | Neurological/-muscular disorders | G1, G20-21, G23-24, G40.5, G61.0, G70-73, F84.0-84.1, Q05.0-05.6, G80.0, G80.2-80.3 | - |
| High | Kidney disease | N18.3-18.5 | - |
| High | Liver disease | K70.4, K72 | - |
| Moderate | Diabetes mellitus | E10-14 | T89-90 |
| Moderate | Lung disease | E84, J41-J47, J84, J98 | R95-96 |
| Moderate | Overweight and obesity | E66 | T82 |
| Moderate | Cardiovascular disease | I05-09, I2, I31-32, I34-37, I39-43, I46, I48-50 | K74-78, K82-83, K87 |
| Moderate | Stroke | I60-64, I69.1-69.4, I69.8, I69.0 | K90-91 |
| Moderate | Dementia | F0, G30-31 | P70 |
| Moderate | Reduced immunity | G35, M05-09, M13-14, K50-51 | - |
