## Supplementary material for "Medical Risk Classification For Severe COVID-19 Based On Chronic Medical Conditions: A Comparative Analysis": Supplementary Table 4.docx

**Supplementary Table 4** Overview of the most prevalent chronic medical condition categories to show the differences between classification selection of ICD-10 and ICPC-2 codes within a category.

|  | Risk group | | General population | | Hospitalized population | | Deceased population | |
| --- | --- | --- | --- | --- | --- | --- | --- | --- |
| Classification | National | European | National | European | National | European | National | European |
| **Prevalence in the Netherlands** |  |  |  |  |  |  |  |  |
| Lung disease | moderate | high | 4.6% | 1.9% | 20.5% | 14.7% | 27.2% | 18.5% |
| Cardiovascular disease | moderate | high | 4.3% | 4.5% | 21.5% | 23.3% | 32.4% | 33.1% |
| Diabetes mellitus | moderate | high | 1.8% | 1.7% | 13.7% | 13.5% | 20.9% | 19.6% |
| Cancer | high | high | 0.5% | 2.0% | 4.1% | 9.1% | 4.1% | 10.8% |
| Kidney disease | moderate | moderate | 2.3% | 1.3% | 15.2% | 11.3% | 23.9% | 18.2% |
| **Prevalence in Norway** | moderate |  |  |  |  |  |  |  |
| Lung disease |  | high | 4.4% | 1.8% | 20.4% | 15.7% | 12.3% | 12.9% |
| Cardiovascular disease | moderate | high | 4.9% | 3.6% | 25.1% | 22.6% | 29.2% | 27.0% |
| Diabetes mellitus | moderate | high | 3.8% | 2.3% | 24.4% | 16.2% | 17.7% | 12.9% |
| Cancer | high | high | 2.3% | 2.1% | 12.7% | 13.6% | 11.7% | 11.3% |
| Kidney disease | high | moderate | 0.2% | 0.8% | 2.4% | 10.1% | 2.3% | 13.3% |
