## Supplementary figures and images for "Medical Risk Classification For Severe COVID-19 Based On Chronic Medical Conditions: A Comparative Analysis"

### Suppl Fig 1.tiff

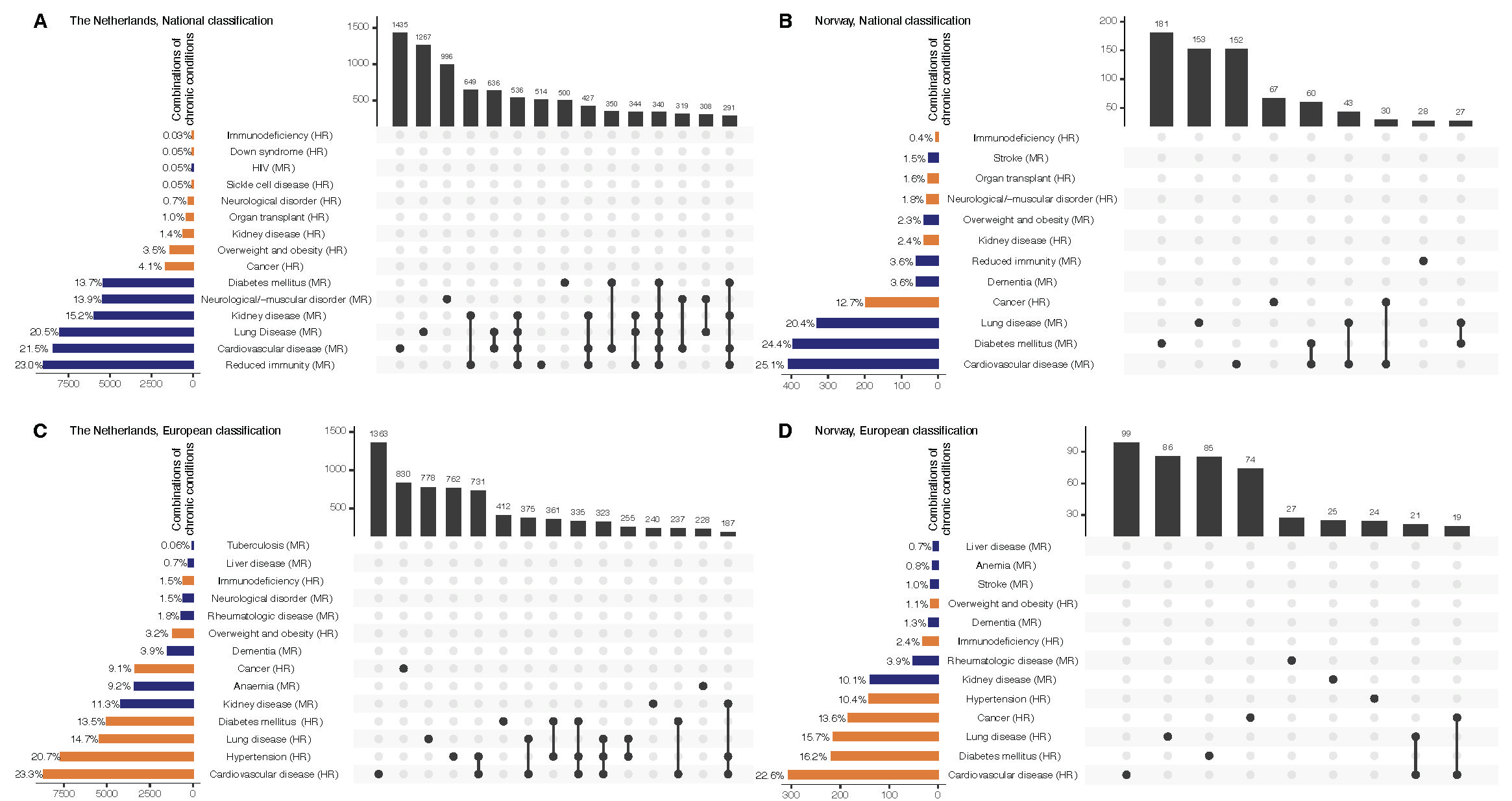

### Suppl Fig 2.tiff

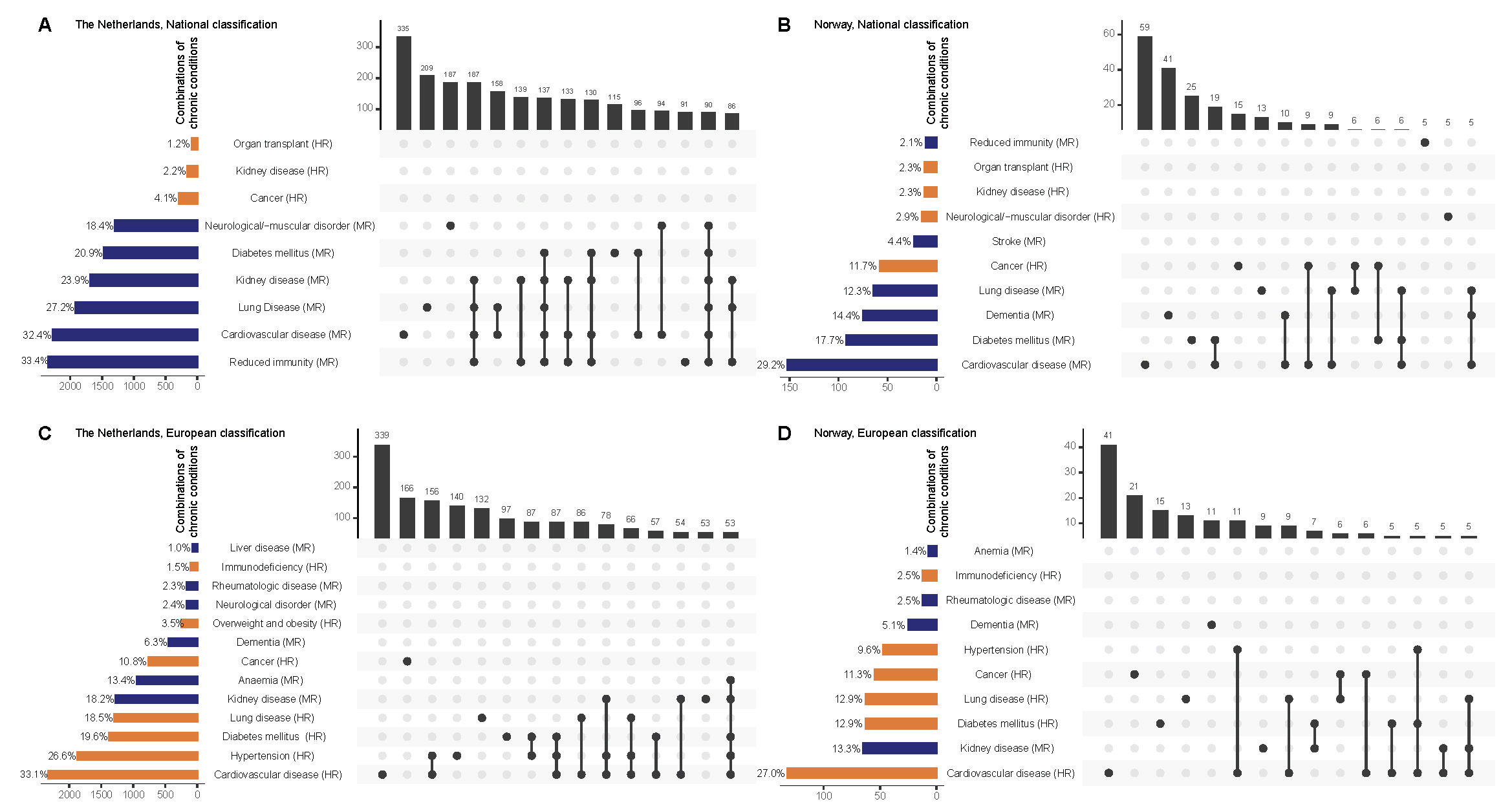

### Suppl Fig 3.tiff

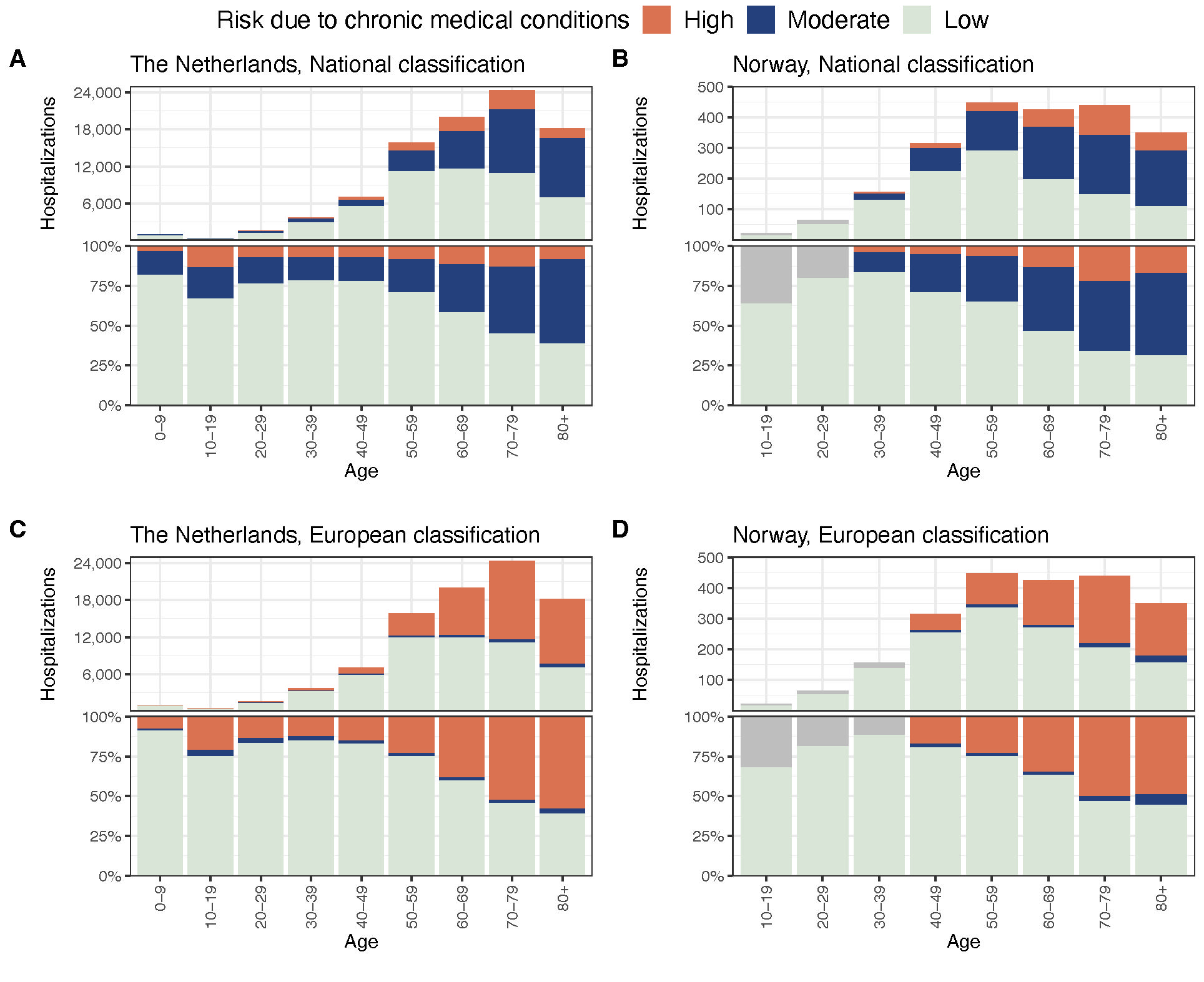

### Suppl Fig 4.tiff

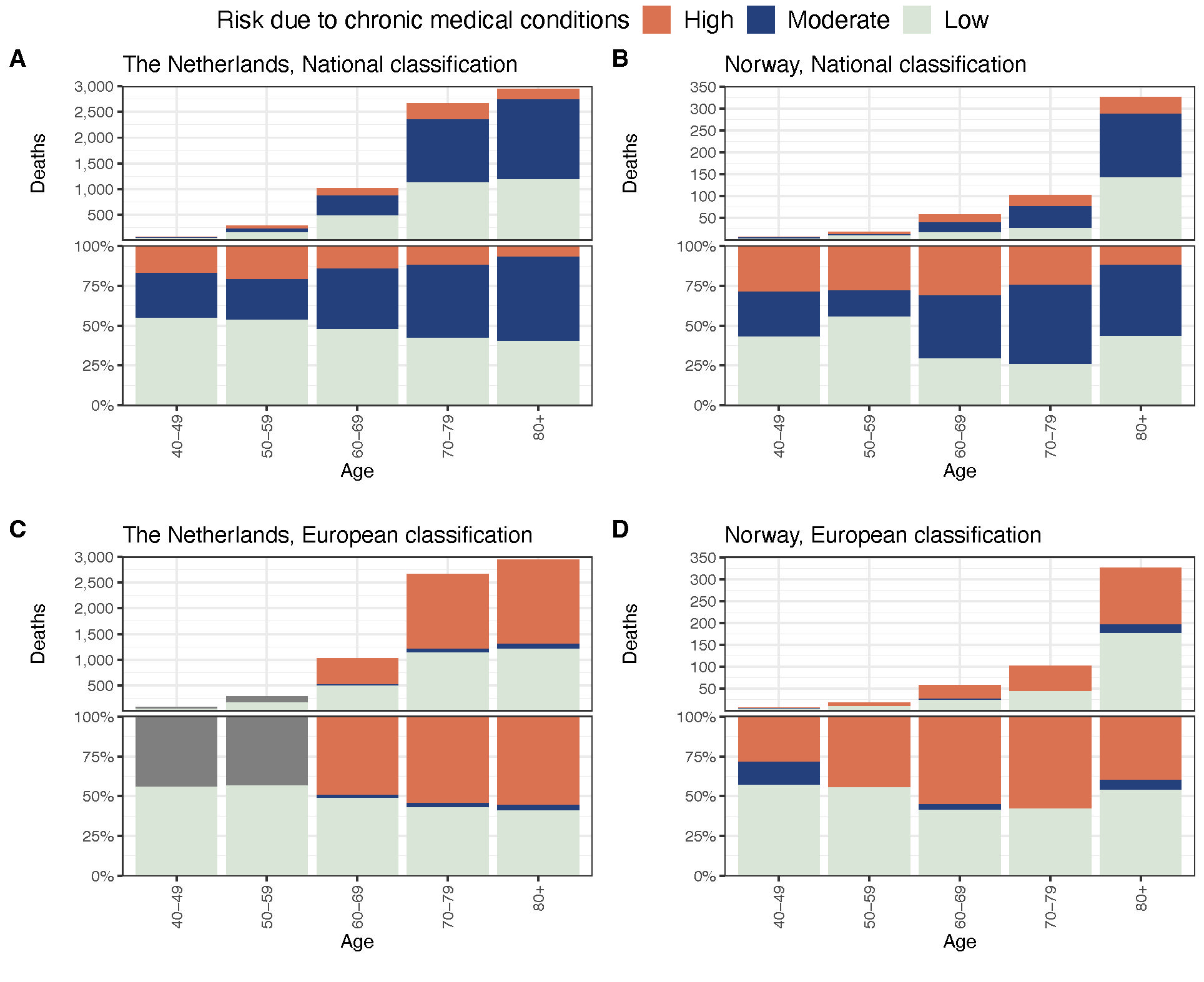

### Suppl Fig 5.tiff

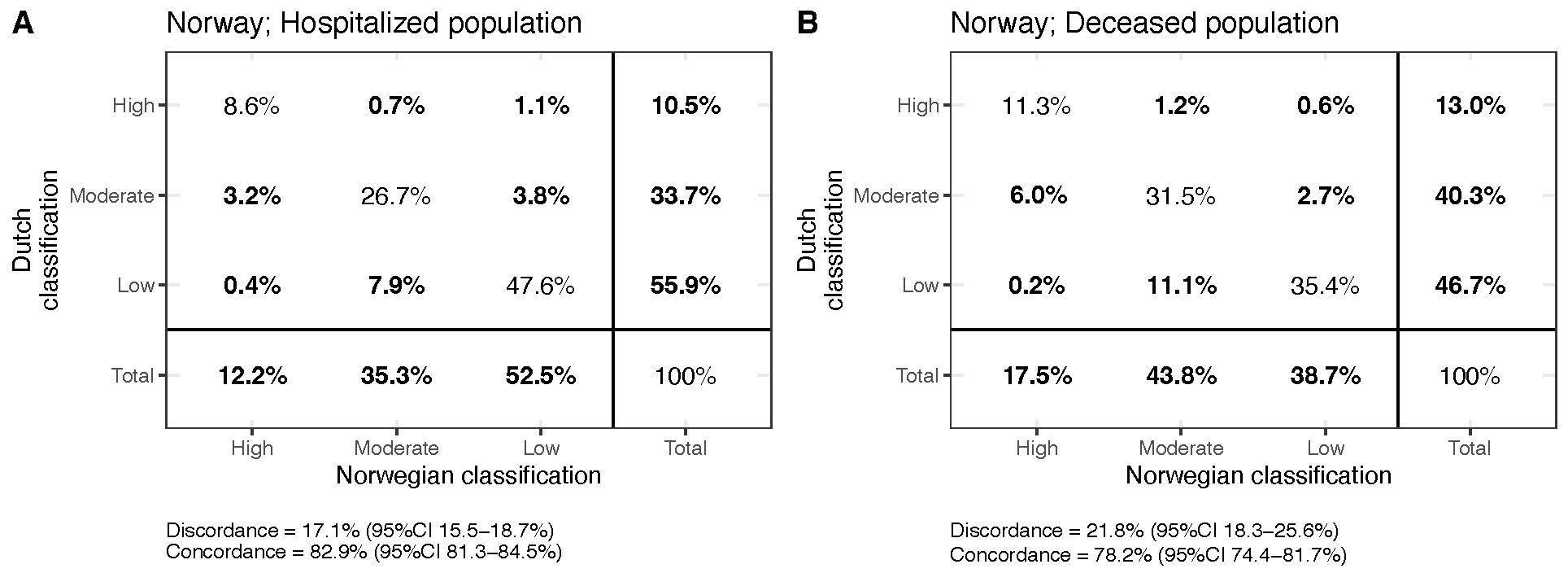

### Suppl Fig 6.tiff

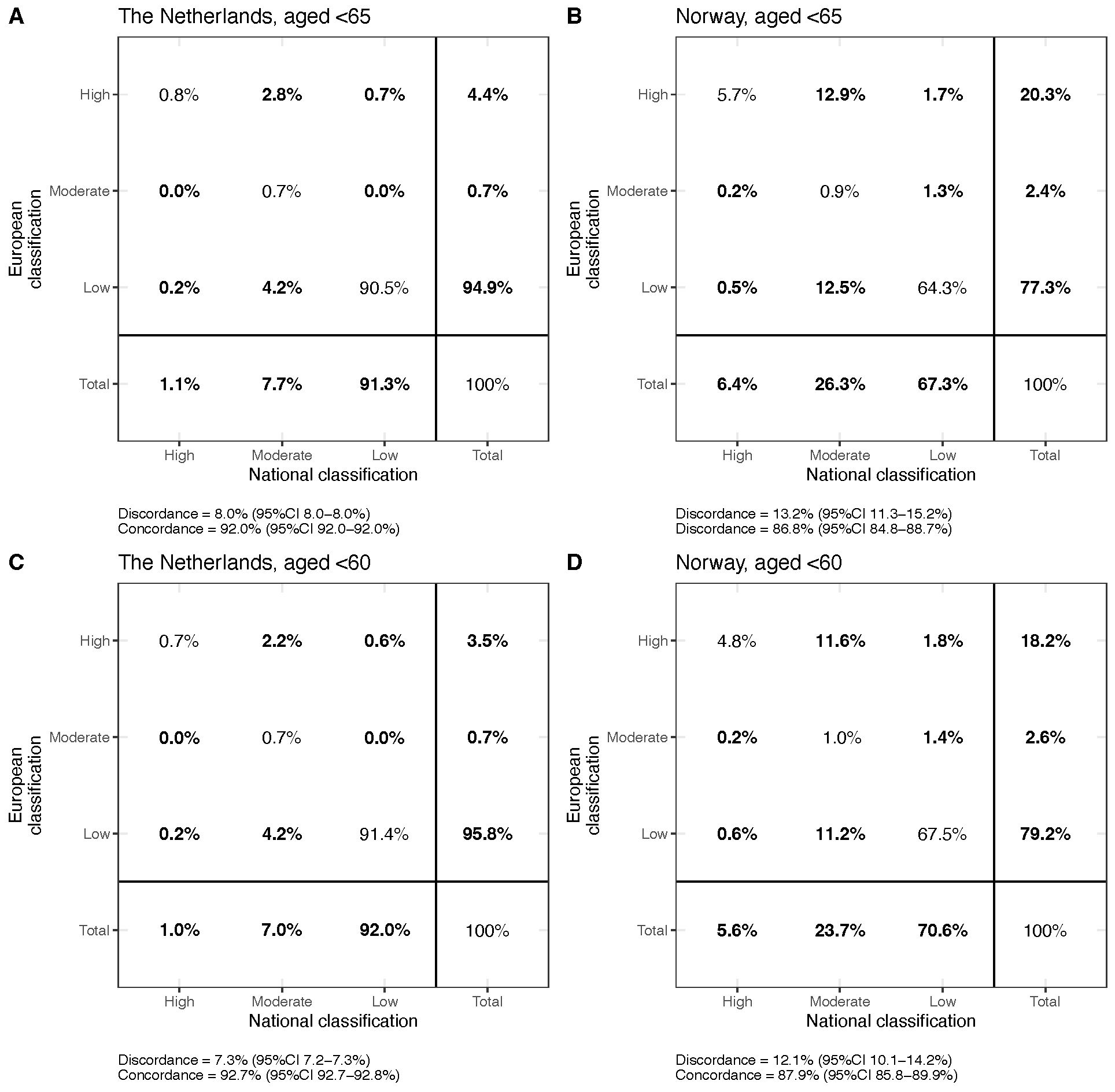

### Suppl Fig 7.tiff

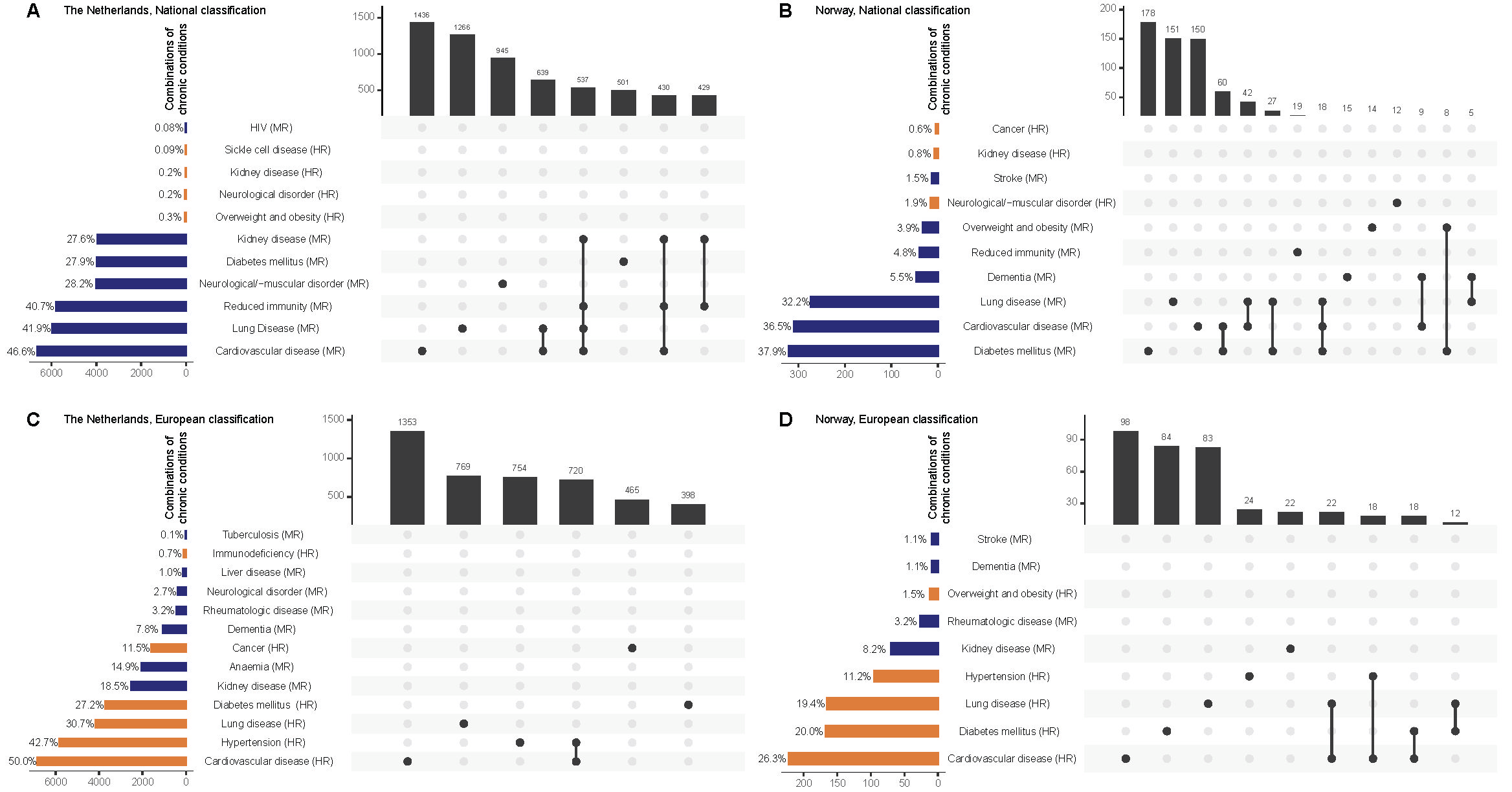

### Suppl Fig 8.tiff

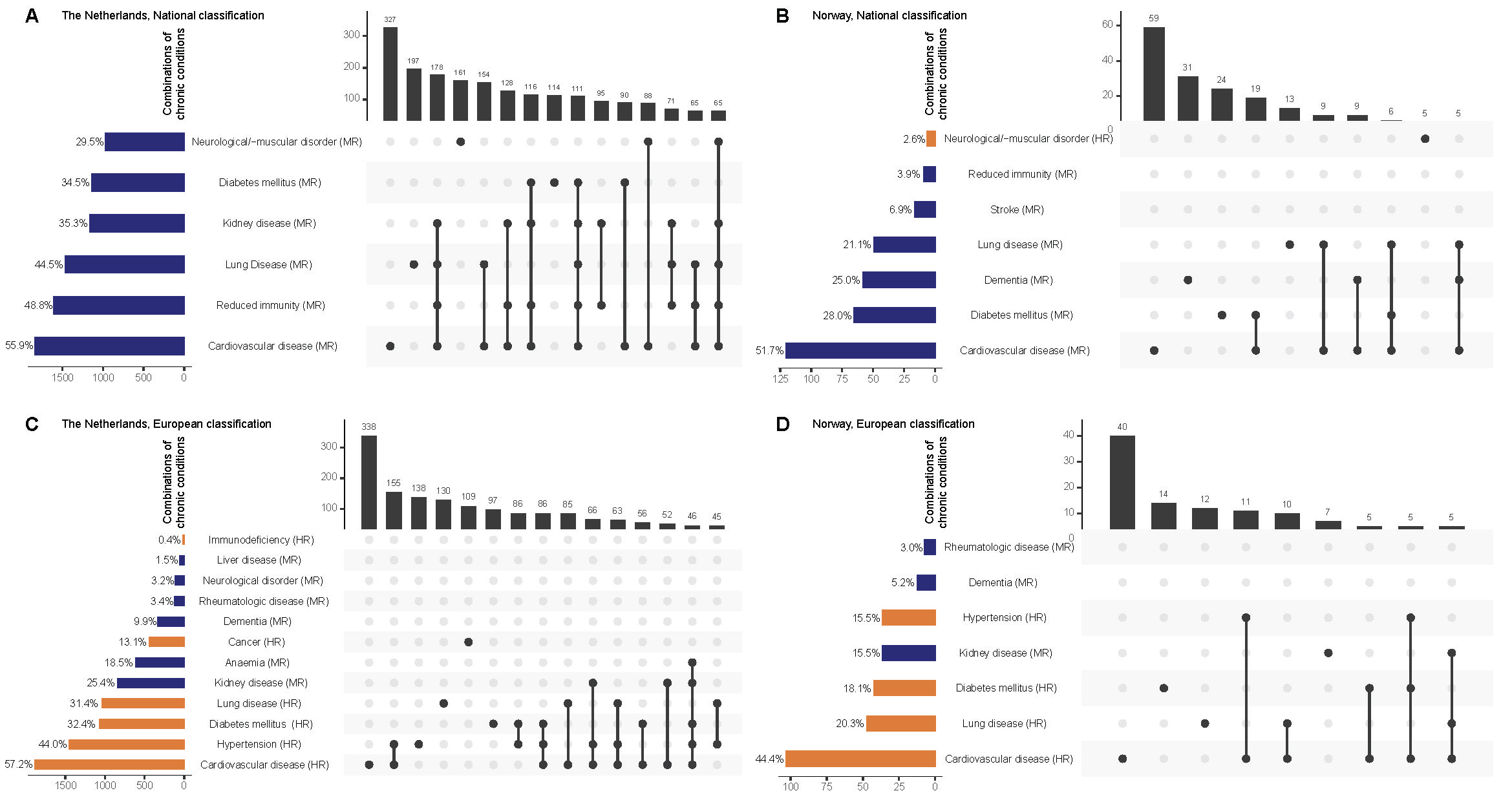

### Suppl Fig 9.tiff

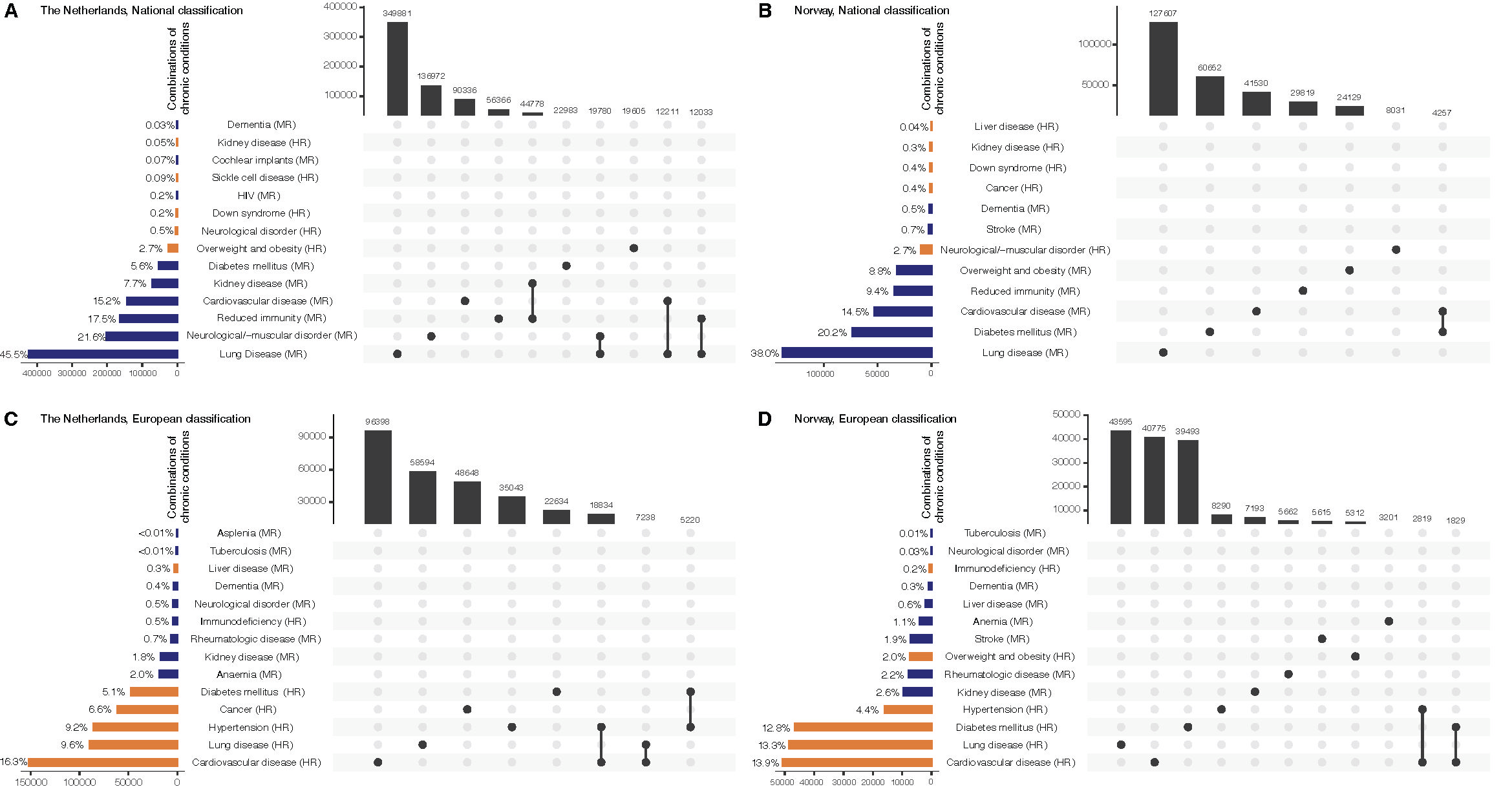

### Suppl Fig 10.tiff

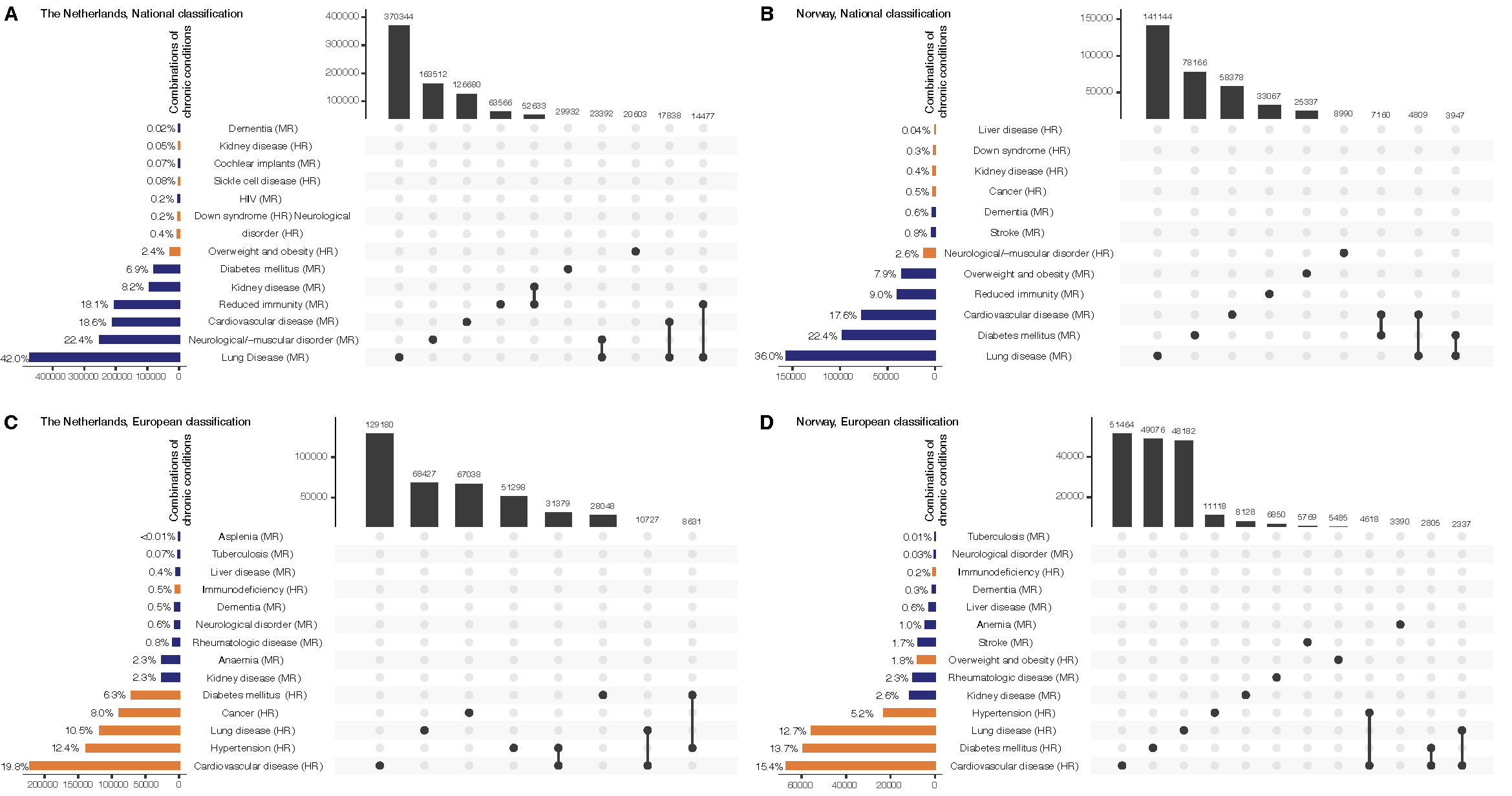
